# A Decade of Radiological Examinations in the Emergency Department: A Monocentric Retrospective Observational Study

**DOI:** 10.1101/2024.12.04.24318507

**Authors:** Ndiaw Goumballa, Fabien de Oliveira, Fabien Coisy, Jean Goupil, Julien Frandon, Florence Longueville, Catherine Daladouire, Romain Genre Grandpierre, Jean Paul Beregi

**Affiliations:** Department of Medical Imaging, Nîmes University Hospital, 30029 Nîmes, France; IMAGINE UR UM 103, Montpellier University, Department of Medical Imaging, Nîmes University Hospital, 30029 Nîmes, France; Department of emergency, Nîmes University Hospital, 30029 Nîmes, France

**Keywords:** Radiology, Emergency, CT scan, MRI, Ultrasound, X-Ray, Imaging, Epidemiology

## Abstract

**Purpose:** To analyze the evolution of various radiological examinations requested for patients admitted to the emergency department (ED) of a University Hospital.

**Materials and Methods:** We analyzed data on MRI, CT scan, Ultrasound (US), and X-ray activities, along with ED patient admissions over a ten-year period from 2014 to 2023. The patient data for each modality, categorized by age and sex, were extracted from the Radiological Information System database.

**Results:** The number of patients undergoing radiological examinations increased from 35,532 in 2014 to 44,592 in 2023, marking a 25.9% increase, while the number of ED admissions rose from 71,776 to 103,456, a 44.1% increase. The ratio of radiological examinations to ED admissions decreased from 49.5% in 2014 to 43.1% in 2023. Over the study period, the number of patients receiving MRI and CT scans increased by 80.4% and 105.8%, respectively. The number of patients undergoing ultrasound remained relatively stable, with 2,616 in 2014 and 2,432 in 2023. In contrast, the number of X-ray patients decreased by 33.1% from 22,236 in 2014 to 14,847 in 2020 but rebounded to 20,492 in 2023. Male patients more frequently underwent CT (51.7%) and X-ray examinations (53.3%).

**Conclusion:** This study highlights a significant increase in radiological activity within the emergency department, especially in CT scans and MRI usage over the ten-year period while ultrasound examinations stayed flat, accompanied by a decline in the ratio of radiological examinations to ED admissions.

## 1. Introduction

Radiological examinations using computed tomography (CT) and magnetic resonance imaging (MRI) have steadily increased in recent years within emergency departments (EDs), in a tertiary care ED in Korea [1]. Between 1993 and 2007 in American hospitals, the use of imaging in emergencies escalated from 276.8 per 1,000 ED visits to 852.8 per 1,000 ED visits [2]. Several studies have indicated that this rise was particularly associated with increased usage of CT and MRI [3]. During the COVID-19 pandemic, a study highlighted the effectiveness of using chest CT in emergency radiology for diagnosing COVID-19 pneumonia in France [4]. Furthermore, in 2015, Bertin et al. noted that radiography (X-ray) was excessively used in EDs, especially for plain abdominal imaging [5]. Additionally, numerous studies have underscored the significance of using ultrasound (US) in managing emergency patients [6,]. However, in France, there is a lack of data on the evolution of radiology examinations in EDs.

EDs serve as a critical nexus between outpatient and inpatient care, reflecting key healthcare environments. Their role is to provide immediate access to care for all patients, serving as a gateway to hospital-based inpatient services without an appointment, regardless of whether the patient’s condition is medically urgent or perceived as such [7]. Managing patients admitted to the ED often involves conducting additional tests to determine diagnoses, including radiology [3].

The aim of this study was to perform a descriptive analysis of the evolution of radiological examination activities (MRI, CT, US, and X-ray) performed on patients admitted to the ED at a French university hospital over a ten-year period (2014 to 2023).

## 2. Materials and methods

### 2.1. Study Setting

This retrospective study was conducted between July 1, 2024 and July 31, 2024, analyzing data from the institutional Radiological Information System (RIS, XPlore Analysis, version 7.0, EDL, Paris, France), covering the period from 2014 to 2023. A RIS is a specialized software system designed to manage medical imaging data and associated operations within radiology departments. Its primary functions include scheduling patient appointments, tracking radiology imaging orders, storing images and reports, and managing billing information. Patients are registered on the RIS according to their origin and nature of demand: inpatient (H), emergency (U), outpatient (E), day care unit (J), etc. Additionally, it facilitates multi-criteria analysis queries across all accessible information within the radiological database. We had access to anonymized information such as age, gender and imaging modality of patients. A non-opposition letter was sent to patients, explaining the purposes of the study and their right to object to the use of their data. This retrospective observational study was approved by the institutional review board.

### 2.2. Data collection

Patients for this study were identified in the Radiological Information System (RIS) database based on their emergency status (U). Subsequently, we restricted our analysis to the period from January 1, 2014 to December 31, 2023. Finally, we included in our filter the following imaging modalities: TDM, MRI, US and X-ray. We collected for all patients, the demographic information (age and sex) and imaging modalities from the RIS. Additionally, data on the number of patients admitted to the emergency department were sourced from the hospital operating system. The total number of patients who underwent imaging examinations was assessed by modality and compared to the total number of emergency room admissions annually. An analysis spanning the ten-year period was conducted. The percentage of increase from one year to the next corresponds to the percentage change rate and was calculated using the following formula: ([new value - old value] / old value) x 100. We calculated a ratio of the number of patients who underwent radiological examinations to the number of patients admitted to the ED.

## 3. Results

### 3.1. Evaluation of imaging by modality for patients in emergency department

The number of patients admitted to emergency departments rose from 71,776 in 2014 to 103,456 in 2023, an increase of 44.1% **(Table 1, Supplementary figure 1)**. However, a significant decrease of 22.9% was observed in 2020, with patient admissions dropping to 77,767 from 95,548 in 2019. Over the same period, the total number of patients receiving radiology examinations increased from 35,532 in 2014 to 44,592 in 2023, marking a 25.9% rise. The ratio of radiological examinations to emergency admissions decreased from 49.5% in 2014 to 43.1% in 2023.

**Table 1:**
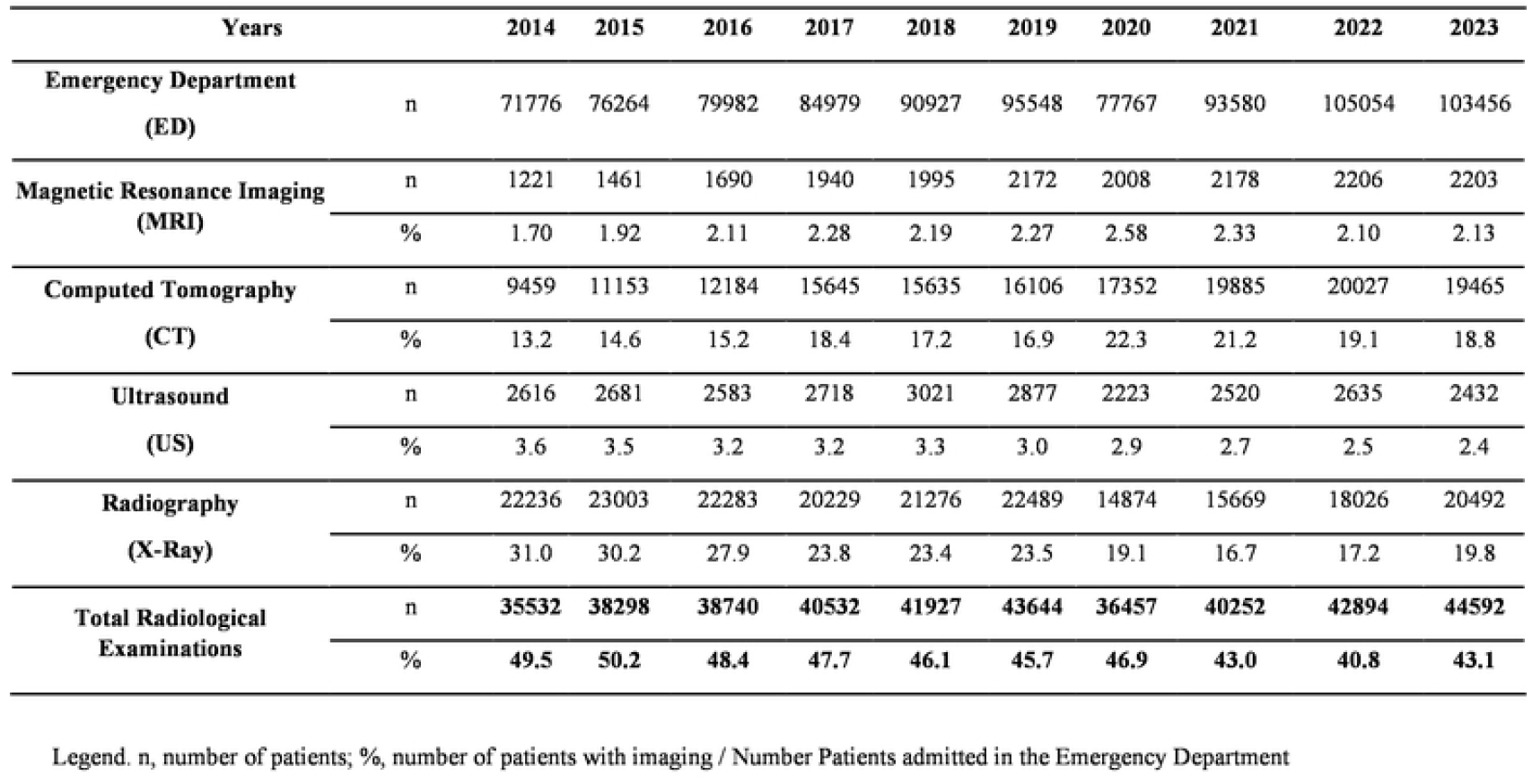
Total Number of Examinations by Modality Performed in Emergency Imaging.

**Figure 1.**
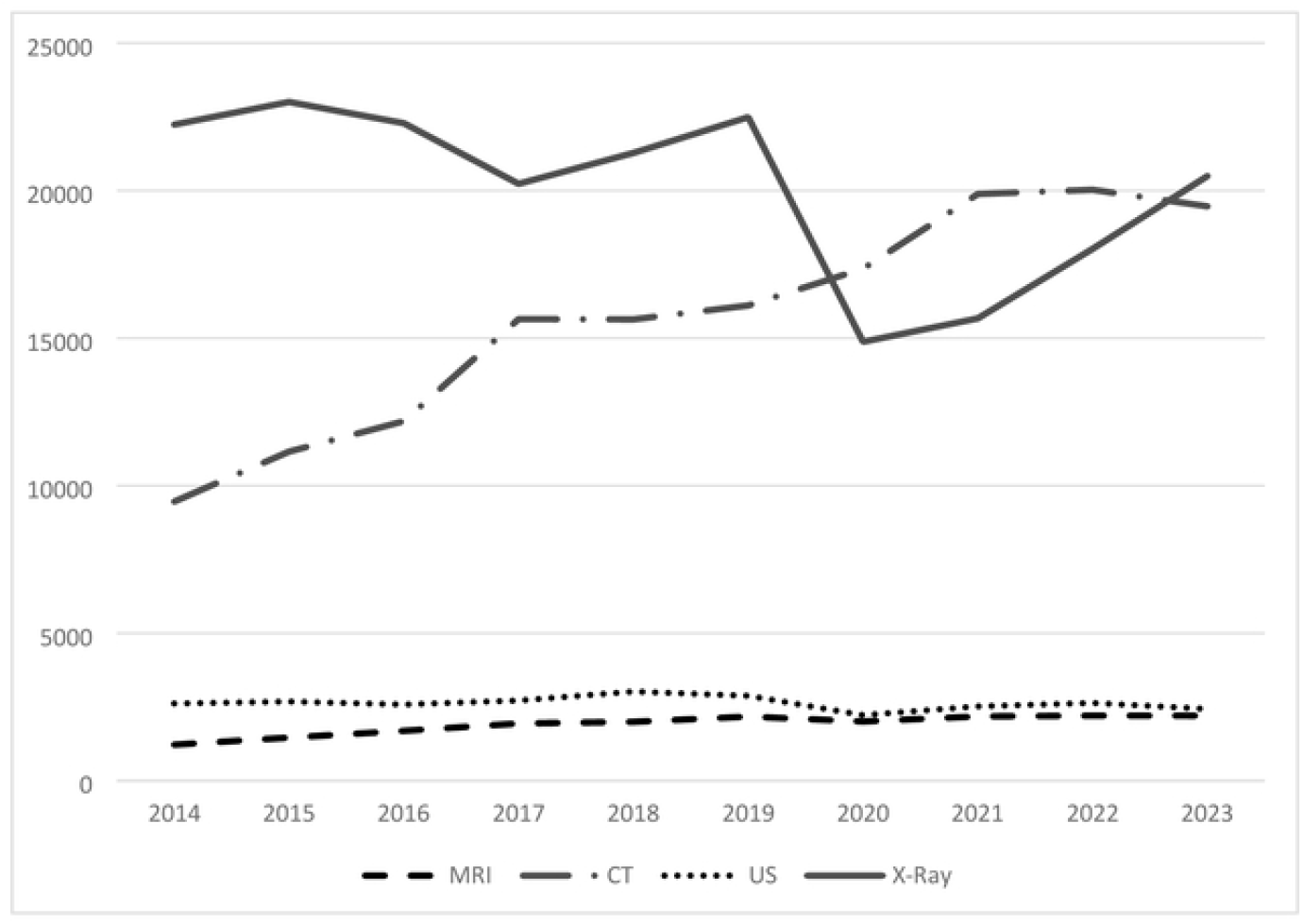
Total Number of Examinations Performed by Modality in Emergency Imaging. Legend. MRI, Magnetic Resonance Imaging; CT, Computed Tomography; Us, Ultrasound; X-Ray, Radiography

The analysis of patient trends over the ten-year period by modality is detailed in **Table 1 and Figure 1**. The number of patients receiving MRI increased by 80.4% from 2014 to 2023 (from 1,221 to 2,203 patients). This growth occurred from 2014 to 2019, after which the numbers plateaued around 2,200 patients, though a slight decrease was noted in 2020. The annual number of CT examinations saw a 111.7% increase from 2014 to 2022 (from 9,459 to 20,027 patients), stabilizing in 2023 at 19,465, reflecting a 105.8% increase from 2014. The annual number of US examinations remained stable throughout the ten-year study, ranging from 2,616 in 2014 to 2,432 in 2023, with a peak of 3,021 in 2018, a net increase of 15.5% compared to 2014.

The number of annual X-ray patients remained stable from 2014 to 2019, ranging from 22,236 to 22,489 patients. A marked decrease of 33.1% occurred in 2020, with only 14,874 patients undergoing X-rays. A slight recovery was noted by 2023, with the numbers increasing to 20,492, although this was still lower than the 2019 figures.

### 3.2. Characteristics of the patients over the ten-year period

During the study period, a total of 19,074 MRI scans, 156,910 CT scans, 26,306 US sessions, and 200,576 X-ray examinations were conducted **(Table 2)**. Males were more likely to undergo CT (51.7%) and X-ray examinations (53.3%), while females were more frequently represented in MRI (52.5%) and US (52.6%) groups. Older patients, aged over 70, were more likely to undergo CT scans (41.8%) and MRIs (37.7%), with mean ages of 58.8 and 58.3 years, respectively **(Table 2)**. In contrast, the US and X-ray modalities were most prevalent among the under 20 age group, accounting for 53.0% and 23.2% of cases, respectively. The average age of patients undergoing US was relatively young at 29.7 years, and for X-rays, it was 47.1 years. Over the course of the study, the average age of patients undergoing CT and MRI increased. However, the average age of patients receiving X-rays decreased from 50 to 41 years between 2014 and 2021. Similarly, there was a gradual decrease in the average age of patients undergoing US, from 36 to 20 years between 2014 and 2023 **(Figure 2)**.

**Table 2:** Patient Characteristics from 2014 to 2023.

|  |  | **MRI**<br>**(n=19,074)** | **CT**<br>**(n=156,910)** | **US**<br>**(n=26,306)** | **X-Ray**<br>**(n=200,576)** |
| --- | --- | --- | --- | --- | --- |
| **Gender** | **Male** | 9,041<br>(47.4) | 81,063<br>(51.7) | 12,457<br>(47.3) | 10,6813<br>(53.3) |
|  | **Female** | 10,023<br>(52.5) | 75,680<br>(48.2) | 13,829<br>(52.6)) | 93,500<br>(46.6) |
|  | **Unspecified** | 10<br>(0.1) | 167<br>(0.1) | 20<br>(0.1) | 263<br>(0.1) |
| **Age (years)** | **Mean** | 58.3±21.9 | 58.8±24.9 | 29.7±28.4) | 47.1±29.0 |
|  | **Min-Max** | [0-103] | [0-108] | [0-105] | [0-114] |
|  | **<20** | 1,155<br>(6.1) | 12,711<br>(8.1) | 13,947<br>(53.0) | 46,550<br>(23.2) |
|  | **[20-29]** | 1,370<br>(7.2) | 14,071<br>(9.0) | 2,584<br>(9.8) | 23,823<br>(11.9) |
|  | **[30-39]** | 1,727<br>(9.0) | 13,026<br>(8.3) | 1,834<br>(7.0) | 18,560<br>(9.2) |
|  | **[40-49]** | 2,033<br>(10.7) | 14,267<br>(9.1) | 1,236<br>(4.7) | 17,069<br>(8.5) |
|  | **[50-59]** | 2,491<br>(13.1) | 17,031<br>(10.8) | 1,184<br>(4.5) | 17,497<br>(8.7) |
|  | **[60-69]** | 3,100<br>(16.2) | 20,264<br>(12.9) | 1,487<br>(5.6) | 18,005<br>(9.0) |
|  | **[70-79]** | 3,658<br>(19.2) | 24,897<br>(15.9) | 1,673<br>(6.4) | 20,184<br>(10.1) |
|  | **≥80** | 3,540<br>(18.5) | 40,643<br>(25.9) | 2,361<br>(9.0) | 38,888<br>(19.4) |

**Figure 2.**
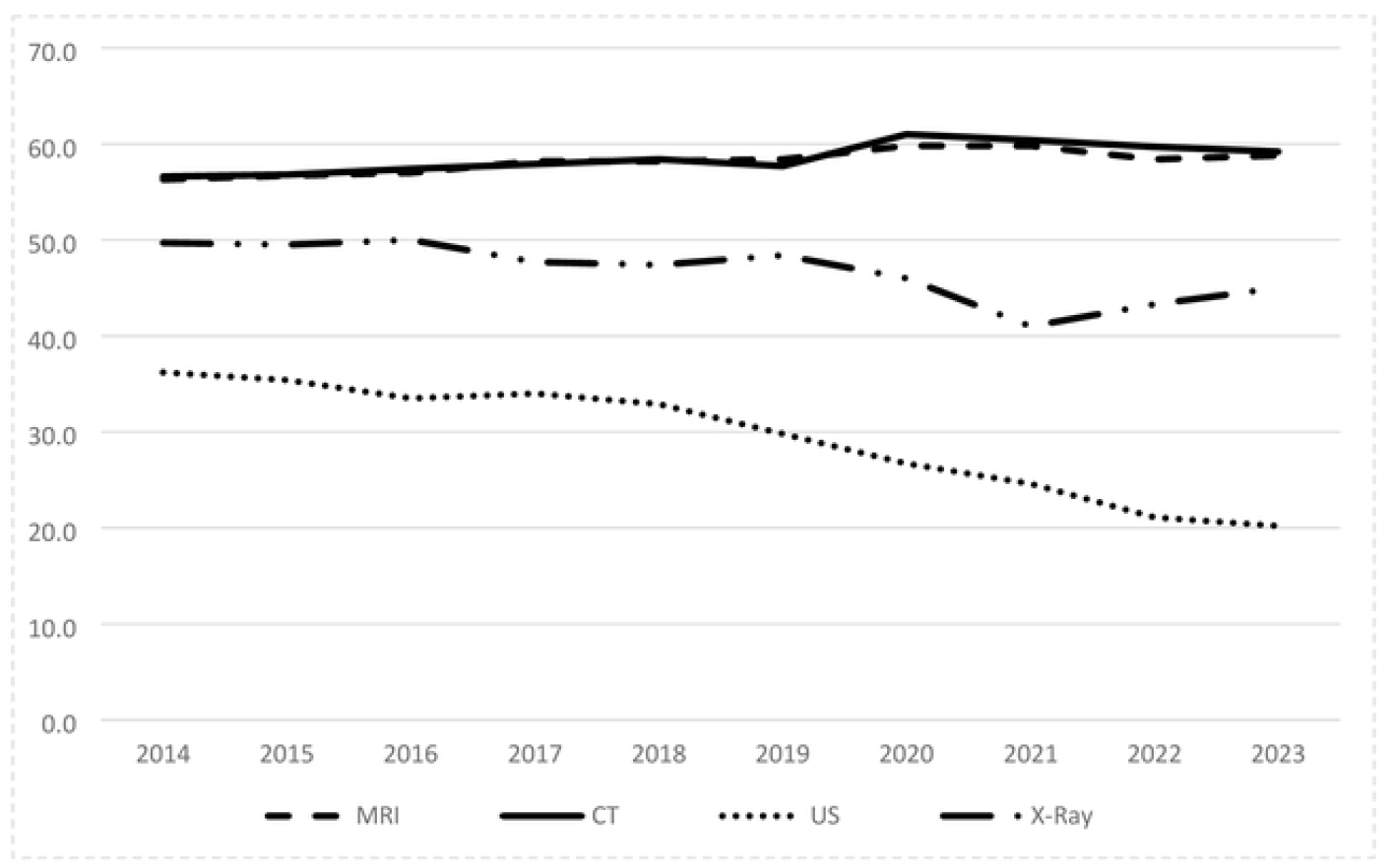
Annual Variation in Average Age by Modality from 2014 to 2023. Legend. MRI. Magnetic Resonance Imaging; CT, Computed Tomography; Us, Ultrasound; X-Ray, Radiography

## 4. Discussion

Over the 10-year study period at our university hospital, the primary finding was that the ratio of radiological examinations to ED admissions decreased from 49.5% in 2014 to 43.1% in 2023, despite an increase in patient intake for both departments.

The number of patients admitted to the Emergency Department (ED) surged by 44.1% while patients referred to radiology from the ED increased by 25.9%, with a significant rise in the use of CT scans. This growth in radiological examinations within the ED aligns with findings from several international studies [1,2]. Our results, showing an increasing utilization of radiological exams in the ED from 2014 to 2023, are consistent with these studies. However, the ratio of examinations to admissions decreased, which can be attributed to multiple factors. One potential reason is the higher admission rates of patients with social or psychological issues who do not require imaging analyses [8]. Another factor could be the severity scores of patients, although these scores were not available in 2014 for patients entering the ED; the severity is influenced by the services offered by public or private institutions in the area. A third factor, could be the use of fast duplex by emergency doctors inducing less examinations. Last factor is the collaboration established in 2013 between the radiology and emergency departments, initiated during a period of significant challenges related to appropriateness.

Since then, monthly meetings between emergency doctors and radiologists, along with frequent training sessions on selecting the optimal exams for the right patients, research, and publications particularly concerning ultra-low dose (ULD) CT scans have been implemented. During these regular meetings, appropriateness and best practices are systematically discussed in light of national and international publications and recommendations.

Our results indicated an increasing utilization of MRI in emergency departments (EDs), particularly among female and older patients. Anh S et al. reported a similar rise in MRI use from 2001 to 2010 [1]. The application of MRI in EDs, particularly for managing appendicitis and brain injuries, has been well documented in several studies [9]. A significant increase in MRI usage during the first seven years was also driven by improved access to MRI facilities in our region, and the leveling off observed in the last three years reflects an adjustment to meet the population’s needs. Additionally, we noted a substantial overall increase in CT scan usage in EDs throughout the study period. This rise could partly be attributed to increased awareness about the benefits of ultra-low-dose CT in our radiology department [10–12]. The number of CT examinations performed increased by 20%, predominantly among older patients, echoing findings from other studies [7,13]. Enhanced access to CT scans became possible in 2016 with the replacement of an X-ray machine with a CT scanner. In contrast, the annual number of US examinations remained stable throughout the study period, with more frequent use observed in younger patients. Over time, US usage was particularly focused on pediatric and gynecological indications. However, X-ray usage in the ED decreased by 33% after 2019 compared to 2014, a decline that may be attributed to several factors, including the COVID-19 pandemic and the associated lockdowns, which also reduced patient admissions to EDs. Additionally, the efficacy of ultra-low-dose CT scans as a replacement for X-rays has been confirmed in multiple studies [10–12].

The development of artificial intelligence tools for emergency radiology, as noted in several studies [14,15], remains crucial. The effective use of imaging in patient management and the triage of examinations to identify pathologies or prioritize radiological assessments will be key areas of focus in the near future to enhance the quality of radiological reports.

Some limitations of this study should be noted. The data were solely sourced from the institutional Radiological Information System (RIS), and there may be instances of missing or incorrect data or data that are difficult to classify. We did not record the reason for patients admissions to the ED as it was not possible to make specific analyses. Given that our study is single-centered, the findings might not represent practices in other emergency departments across France, and may not be applicable in other settings. To obtain comparative insights, it would be beneficial to conduct an annual multicenter study throughout France or to conduct an international survey.

In conclusion, the increase in patient admissions to the emergency department is negatively correlated with the increase in radiological examinations. The reasons for this difference should be investigated through further multicenter studies with population-based criteria.

## Data Availability

Data is available upon request after acceptance.

## Funding

This research did not receive any specific grant from funding agencies in the public, commercial, or not-for-profit sectors.

## Declaration of competing interest

No conflict of interest to declare.

## Author contributions

NG, CD and JPB, conception and experiments of the study. NG and CD collected and analyzed the data. NG and JPB drafted the manuscript. FO, SB, JG, JF, FL and RGG critically reviewed the manuscript. All authors contributed to and approved the current version of the manuscript.

## Notes

### Competing Interest Statement

The authors have declared no competing interest.

### Funding Statement

The author(s) received no specific funding for this work.

### Author Declarations

This study was approved by the Institutional Review Board of CHU Nimes. on the number of 24.06.05.

